# Longitudinal effects of ambient AI scribe use on documentation burden and financial productivity in primary care: A quasi-experimental study

**DOI:** 10.64898/2026.01.12.26343538

**Authors:** RJ Waken, Sunny S. Lou, Mackenzie Hofford, Elise Eiden, Carrie Burk, Seunghwan Kim, Jared Esker, Linying Zhang, Thomas M. Maddox, Joanna Abraham, Albert M. Lai, Sam Bhayani, Deborah O’Dell, Jennifer Schmidt, Kayla Paynter, Michele Thomas, Matthew Gerling, Philip R.O. Payne, Thomas Kannampallil

**Author notes:** Corresponding Author*: Thomas Kannampallil, PhD, Washington University School of Medicine (WashU Medicine), 660 South Euclid Ave, Campus Box 8054, St Louis, Missouri, USA 63110.

## Abstract

**Objective:** To investigate the longitudinal effects of an ambient AI scribe use in primary care on outcomes related to documentation time, note writing patterns, and clinician financial productivity.

**Method:** We conducted a quasi-experimental study with an interrupted time-series design to ascertain the effect of AI scribe use on note writing time, time outside of scheduled hours (TOSH), pajama time (530PM–7AM), note length, note closure <24hours, and work Relative Value Units (wRVU). For each primary care clinician (physician, advanced practice provider), date of initial access was day zero; each measure was calculated for 150 days prior (pre-period) and after (post-period) day zero.

**Results:** 220 clinicians (Mean age=43.7, 70.9% females; 56.4% physicians) from 36 clinics, conducting 314,845 patient encounters were included. Compared to the pre-period, there was 7% decrease in note writing time at day zero (Incidence Rate Ratio (IRR) 0.93,95% Credible Interval (CI) [0.89,0.98]), increasing to 16% decrease by day 150 (IRR 0.84,95%CI [0.82,0.87]). There was no change in pajama time, TOSH, or wRVU at day zero; however, at day 150, there was a 19% decrease in pajama time (0.81,95%CI [0.73,0.90]), 13% decrease in TOSH (0.87,95%CI [0.77,0.98]), and a 2% increase in wRVU (1.02,95%CI [1.01,1.04]). At day zero, there was a 5% increase in note closures (1.05,95%CI [1.02,1.08]), with no difference to pre-period levels by day 150. There were no changes in note length at day zero or day 150.

**Discussion:** There were persistent changes in AI scribe use measures over time as clinicians situated these tools within their workflows.

## Introduction

The burden of electronic health record (EHR) use has increased exponentially over the past decade.^1,2^ A primary contributor to this burden is the increased documentation requirements, owing to policy and payment requirements, increasing clinician workload, burnout, and work-life balance.^3–5^ Research has highlighted that every hour of patient-facing time was associated with ~2 hours of additional documentation, potentially detracting clinicians from direct patient interactions, affecting both patient care, and clinician satisfaction.^6^

Artificial Intelligence (AI)-based scribes (henceforth, “AI scribes”), have the potential to reduce clinician time spent on documentation, by generating the draft of a clinical note from the patient-clinician conversation.^7^ These tools have transformed clinical note generation and organization into a language-optimized, model-driven report, with the clinician playing the role of an editor.^8^ Although AI scribes have been rapidly adopted and several studies have been conducted, confirmatory evidence regarding their benefits are limited.^7,9^ Among the reported studies, there are contrasting findings related to savings in documentation time associated with the use of AI scribes. For example, although some studies have reported savings in note writing time after the implementation of an AI scribe (e.g.,^9–12^), others have reported no or marginal time savings (e.g.,^13,14^). Additionally, almost all studies using self-reported survey-based measures have reported on increased clinician satisfaction, reduced burnout, reduced task load, and perceived savings in note writing time.^11,15–19^

These contrasting findings can be attributed to several methodological and measurement considerations in prior studies. First, measurement mechanisms for time-based reporting has relied on vendor-provided metrics, aggregated at a clinician-level, often at bi-weekly or at monthly intervals (e.g., using Epic’s Signal platform^9,13^). Such an aggregation does not capture patient encounter-level variability (e.g., documentation time associated with a specific encounter such as an annual wellness visit) and precludes the ability to control for important encounter-level covariates (e.g., evaluation and management codes associated with a visit) or physician productivity associated with a specific encounter (e.g., work relative value units, wRVU). The lack of encounter-level measures also affects the ability to assess the impact of AI scribes on downstream quality or safety outcomes. These aggregated approaches have also been used for recent randomized controlled trials of AI scribes^13,20^ that have reported limited productivity gains, as highlighted in an accompanying editorial that claimed that AI scribes are not “productivity tools yet.”^21^ In their editorial, Kim et al emphasized that the marginal effect sizes on productivity gains may be potentially related to the aggregated measurement approaches associated with the use of vendor-developed platforms used in these trials.^21^

Second, studies have almost exclusively relied on pre-post study designs, with measurements at two time-points (e.g., 1-month before/after implementation; see exceptions^13,20^). However, as with any new technology, AI scribe adoption and adaptation evolves over time; as such, single time-point measurements are often unable to capture user adaptations that evolve over time.^22–24^

The current study investigates temporal patterns of encounter-level clinician documentation metrics using a causal inference framework, which allows for the investigation of how AI scribes change physician work patterns, and how those changes in work patterns evolve over time. Towards this end, we conducted a longitudinal, quasi-experimental study in primary care to ascertain the impact of an AI scribe on outcomes related to documentation time (note writing time, time outside of scheduled hours, pajama time), note writing patterns (note length, note closure) and clinician financial productivity (wRVUs) at approximately monthly time points over a 150-day period (e.g., 0-, 30-, 120- and 150-days).

## Methods

### Study setting, design, and participants

A longitudinal quasi-experimental study was carried out across outpatient clinics at a large midwestern healthcare system. Primary care clinicians (attending physicians and/or advanced practice providers (APPs)) were provisioned access to an ambient AI scribe (Abridge Inc, New York) integrated within the Epic EHR (Verona, WI) at different time points between June 19, 2024, and July 30, 2025. The choice of primary care was settings was because it provided a homogenous sample of clinical encounters. Moreover, the use of the AI scribe in specialty practice was sporadic and limited.

For provisioning access, clinicians had to complete a training module that included a short video demonstration of the AI scribe workflow, consent process, and tool set up. Clinicians who required additional support were provided 1:1 virtual consultation with an information systems analyst with experience in setting up and using the AI scribe. In addition, given that multiple clinics were involved, there were AI scribe “roadshows” where a training group set up hands-on demonstrations. Additional support was available to all clinicians in an on-demand format, where they could request a consultation with an information systems consultant at any point.

Primary care provider (PCP) encounters, including annual wellness visits, of clinicians provisioned AI scribe access, occurring between January 5, 2024, and October 31, 2025, were included. Clinicians included attending physicians (either MD or DO) or advanced practice providers (APP; NPs or PAs). Patient encounters were excluded if they contained indicators for non-primary care clinicians, telemedicine encounters, and weekend encounters (see STROBE Figure 1 for exclusions).

We used an interrupted time series design to investigate the changes in provider-patient encounter level outcomes after AI scribe tool introduction over a 48-week period (24 weeks pre-implementation, 24 weeks post-implementation) centered on first date of a clinician’s AI scribe access. In this design all clinicians were considered treated as of the day they were first provisioned access (“day zero”), under an intent-to-treat (ITT) framework, while accounting for typical day-to-day variability in clinician work patterns.

The study protocol was approved by the Institutional Review Board of the Washington University School of Medicine (IRB#202511025).

### Data

Data were retrieved from Epic EHR’s User Action Log Lite tables in the Clarity database. These tables record pre-specified clinician activities (e.g., note writing, review, orders), time spent on these activities. Using encounter-related information, we identified specific patient-level details and filtered to primary care encounters only.

For each included patient encounter, we retrieved encounter ID, encounter type (face-to-face, telemedicine, phone), encounter clinician, scheduled appointment hours, whether an AI scribe was used or not (i.e., in the post-period), time spent on note writing activities both during and after the scheduled encounter, note characteristics including length of the note and note closure time, and the billed wRVUs associated with the encounter. In addition, we also captured to the sex and age of both the clinician and the patient associated with an encounter. All data and outcomes were captured at the patient-encounter level; if an encounter was associated with multiple notes, documentation characteristics such as note writing time or character length was aggregated across all included notes, and note closure was defined as the moment that the last encounter note was finalized.

### Outcomes and covariates

There were six outcomes of interest (see Appendix Table 1).

**Table 1.** Summary of clinicians, encounters and outcomes.

|  | **Pre**<br>**(N=138252)** | **Post**<br>**(N=176593)** |
| --- | --- | --- |
| **Clinician specialty** | **Clinicians,**<br>**Encounters** | **Clinicians,**<br>**Encounters** |
| Family Medicine | 109, 73359 | 112, 96017 |
| Internal Medicine | 72, 57576 | 75, 72428 |
| Obstetrics and Gynecology | 22, 298 | 21, 328 |
| Pediatrics | 6, 7019 | 6, 7820 |
| **Clinician credentials** | **Clinicians,**<br>**Encounters** | **Clinicians,**<br>**Encounters** |
| APP | 86, 31618 | 90, 42459 |
| MD/DO | 123, 106634 | 124, 134134 |
| **Abridge Scribe Usage (%)** | NA | 66.7% |
| **Outcomes** | **Mean (SD)** | **Mean (SD)** |
| Note writing time (Mins) | 4.39 (3.78) | 3.83 (3.45) |
| Pajama time (Mins) | 1.71 (6.31) | 1.62 (6.23) |
| Time Outside of Scheduled Hours (Mins) | 1.23 (5.54) | 1.19 (5.79) |
| Note length (Characters) | 8900 (8050) | 9140 (7480) |
| Note closed <24h | 116423 (84.2%) | 148911 (84.3%) |
| Total work Relative Value Units | 2.12 (0.881) | 2.16 (0.856) |

Note writing time was measured as an estimate of the active actions performed on the note including keystrokes, mouse clicks, and mouse movements (e.g., scrolling) with pre-determined timeouts when no activity is detected. Other outcomes included: time outside of scheduled hours (i.e., note writing time outside of scheduled clinic hours, but during a normal workday, 730AM to 5PM); “pajama time” (i.e., time spent on note writing from 530PM to 7AM); note length was number of characters associated with the finalized note; notes signed within the first 24h after the scheduled encounter (note closure <24h); and total work RVUs associated with an encounter as reported in the final billing data.

### Statistical analysis

To estimate the causal effect of the AI scribe introduction on each of the outcomes of interest, we implemented an interrupted time series design. We fitted a Bayesian non-linear interrupted time series model with a latent autoregressive term accounting for day-to-day practice serial variability in clinician work patterns, and modeled non-linear deviations from that using a generalized additive model specification in the post-period using a generalized linear modeling framework.^25^ The latent time series component used the pre-period to tune parameters describing typical serial variability, and in the post-period, accounting for typical serial variability and provided an underlying nowcasted counterfactual, representing the potential untreated outcome; the non-linear intervention effect captured the deviation from the existing serial process, which when combined yields the effect associated with the intervention of interest under the potential outcomes framework. Details of the model including model specification, likelihood, prior parameterization, model diagnostic assessment, model predictions, and spline knot specification, are provided in the Supplementary material (Section B).

Two outputs were generated from the interrupted time series model: (a) the average time-varying change from pre-AI scribe period as characterized by the non-linear intervention effect specification represented as incidence rate ratios in the count response processes and rate ratios in the wRVU process (referred to as IRR hereafter), and (b) the difference in predicted outcomes between the modeled process and counterfactual draws resulting from the posterior predictive distribution aggregated across the 7 day periods starting at day 0, 30, 60, 90, 120, and 150; this prediction incorporated patient encounter-level prediction error resulting in wider interval estimates and predicts the effect of the AI scribe introduction to a new clinician at day 0 (referred to as “predicted changes” at day 0, day 30, day 60, day 90, day 120 and day 150 in the results).

Given the focus on descriptive statistical inference over time in the treatment period, we focus on reporting uncertainty via symmetric 95% credible intervals (CIs) (as this was based on Bayesian inferencing) based on Markov Chain Monte Carlo draws from posterior simulations across four chains rather than hypothesis testing using p-values. We descriptively characterized modeled results as significant, when CI bounds rounded to two decimal places, did not contain 1.

In sensitivity analyses, we also added secular components that adjust for clinician age group at baseline (<36, 36-55, >55) and sex, and for clinician role (physician vs. APP), and we conducted stratified analyses by clinician role (Supplementary Material Section E).

All models were fit using R version 4.6.0 and Stan version 2.36.0, and all preliminary fits to assess our thin plate spline modeling choices were fit using the mgcv package.^26–28^

## Results

### General characteristics

220 clinicians that used AI scribes for any encounter (56% physicians) who worked in 36 clinics were included (Mean age=43.7, 70.9% females) contributed to the sample; n=6 clinicians contributed only to the pre-period and n=11 only to the post period. These clinicians were involved in 314,845 patient encounters during the 48-week time-period centered around their first date of AI scribe access (see Figure 1 for STROBE). Table 1 provides encounter-level pre- and post-AI scribe period summary statistics.

Based on unadjusted analysis, in the post AI scribe period (56.1% of total encounters and 66.7% in which the AI scribe was used), the average per encounter note writing time [4.39 mins (SD=3.78) vs. 3.83 mins (SD=3.45)], TOSH [1.23 mins (SD=5.54) vs. 1.19 mins (SD=5.79)] and pajama time [1.71 mins (SD=6.31) vs. 1.62 mins (SD=6.23)] were marginally lower, compared to the pre-period. However, in the post-period, average total encounter note length [8900 characters (SD=8050) vs. 9140 characters (SD=7480)], average total encounter wRVU [2.12 (SD=0.88) vs. 2.16 (SD=0.85)], as well as the percentage encounters with all notes closed within the first 24 hours [84.2% vs. 84.3%] were higher.

Time series for each of the outcomes over the 48-week period are shown in (Supplementary Material (Section C) and are represented as the average response per encounter at the day-level, along with a time zero marker separating the pre- and post-AI scribe periods.

### Interrupted time series of AI scribe effects

Figure 2 illustrates the post-AI scribe initiation IRRs for each outcome over time with 95% CIs; these represent the average change after AI scribe introduction per encounter after accounting for time varying trends. Table 2 provides a summary of the IRR and predicted changes in outcomes in the clinician week starting at day 0 and at day 150 in the post period for a new clinician introduced to the AI scribe.

**Table 2.** Incidence Rate Ratios (IRR) and predicted changes at day 0 and day 150. *Indicates significant IRRs.

| Outcome measure | IRR at Day 0 | Predicted Change at Day 0 |  | IRR at Day 150 | Predicted Change at Day 150 |
| --- | --- | --- | --- | --- | --- |
| Note writing time (mins) | 0.93, (0.89, 0.98)* | 32.04, (8.95, 54.74)* |  | 0.84, (0.82, 0.87)* | 69.67, (51.8, 87.9)* |
| Pajama time (mins) | 0.95, (0.79, 1.12) | 7.35, (-23.91, 41.18) |  | 0.81, (0.73, 0.90)* | 35.61, (7.4, 65.86)* |
| Time Outside of Scheduled Hours (mins) | 0.89, (0.74, 1.05) | 13.34, (-12.87, 43.62) |  | 0.87, (0.77, 0.98)* | 15.7, (-6.82, 39.3) |
| Note length (1000 Characters) | 1.04, (1.00, 1.09) | -0.08, (-3.87, 4.03) |  | 1.02, (0.99, 1.05) | 0.07, (-4.07, 4.09) |
| Note closed <24h | 1.05, (1.02, 1.08)* | 4.29, (-17.86, 25.71) |  | 0.99, (0.97, 1.00) | -1.43, (-22.14, 20.71) |
| Total work Relative Value Units (wRVU) | 1.02, (1.00, 1.04) | 4.11, (-46.27, 53.15) |  | 1.02, (1.01, 1.04)* | 5.97, (-42.09, 56.17) |

**Note:** IRRs represent the typical trend with 95% CIs, while predicted changes with 95% CIs represent the expected change for a new clinician week and incorporates forecast uncertainty, resulting in wider interval estimates.

At day zero, compared to the pre-period, there was evidence of an immediate 7% decrease on average in note writing time (IRR 0.93, 95%CI [0.89, 0.98]), which continued decrease to 16% below expected time varying trends prior to the AI scribe introduction (IRR 0.84, 95%CI [0.82, 0.87]) by day 150. This translated to approximately 32 minutes of note writing time savings per week at day zero (32.04, 95%CI [8.95, 54.74]) and approximately 70 minutes of note writing time savings per week at day 150 (69.67, 95%CI [51.80, 87.90]).

Compared to the pre-period, there was no evidence of a change in pajama time at day zero (0.95, 95%CI [0.79, 1.12]); however, at day 150 we found evidence of a 19% decrease in pajama time (0.81, 95%CI [0.73, 0.90]). This translated to approximately 35 minutes of pajama time savings per week at day 150 (35.61, 95%CI [7.40, 65.86]).

There was no evidence of a change in TOSH at day zero (0.89, 95%CI [0.74, 1.05]); however, at day 150 we found evidence of a 13% decrease in TOSH (0.87, 95%CI [0.77, 0.98]), compared to the pre-period. This translated to approximately 15 minutes of TOSH savings per week at day 150 (15.70, 95%CI [−6.82, 39.30]).

There was no evidence of increase in note length at day zero (1.04, 95%CI [1.00, 1.09]) or at day 150 (1.02, 95%CI [0.99, 1.05]). At day zero, 5% more notes were closed within 24 hours (note closure <24h 1.05, 95%CI [1.02, 1.08]). This was equivalent to approximately 4.29 additional note closures per week at day zero (4.29, 95%CI [−17.86, 25.71]). By day 150, there was no evidence of differences in the rate of note closures after 24h compared to the pre-period (0.99, 95%CI [0.97, 1.00]).

Finally, there was evidence of a 2% increase total wRVU at day 150 (1.02, 95%CI [1.01, 1.04]). This was equivalent to an additional 5.97 wRVUs per week (5.97, 95%CI [−42.09, 56.17]). Additional IRRs and predicted changes at day 30, 60, 90, and 120 are provided in Supplementary material (Section D).

Additional details of the sensitivity analysis by age, sex and clinician type (physician vs. APP) are provided in Supplementary material (Section E).

## Discussion

Using a longitudinal, quasi-experimental study, we found that the use of an AI scribe decreased note writing time by 15% (~70 minutes per week of predicted time savings) and pajama time by 18% (>30 minutes per week) by day 150. The gain on the clinician productivity was marginal (2% increase per week) by day 150.

During the AI scribe period, there were changes from day zero to day 150 for all considered outcomes, highlighting the complexities of adoption and use of novel AI tools. Longitudinally, changes in outcomes were gradual, but persistent – sometimes changing directions – underscoring the gradual process of adaptation of AI tools, as clinicians situate the tools within their workflows. The changes over time highlight the importance of the longitudinal approach towards studying clinical AI applications.

In terms of the outcomes, several interesting patterns emerged. At day zero, there was a predicted time savings of approximately half-hour per week, and by day 150, the savings were over an hour per week. In comparison, Afshar et al, in their recent RCT, reported note writing time savings of ~22 minutes per day.^20^ In another recent multi-center study, Rotenstein et al reported documentation time savings of ~16 minutes per eight patient scheduled hours.^29^ Our estimate of approximately an hour per week of time savings is lower than the estimates from these studies. There are likely several potential reasons: first, as opposed to much of the prior studies that have relied on vendor-provided metrics (e.g., Epic Signal), we created action-level metrics from audit logs relying on custom data pipelines and data processing. As we have reported elsewhere,^30^ these custom measures are likely underestimates of actual time spent because they rely on granular action events on the EHR (e.g., keystrokes and mouse movement activity). Second, vendor-provided tools, such as Epic’s Signal, aggregate measures at the clinician-level, limiting the ability to directly assess encounter-level outcomes (e.g., RVU or note length per encounter). The current analysis was conducted using encounter-level measures and their longitudinal changes over time (per clinician, per encounter), potentially providing a more accurate, albeit lower estimates of time savings.

Although the wRVU gains were marginal (2% or a predicted 4.4 additional RVUs per week) by day 150, there were differences between physicians and APPs (Supplement, Section E). For example, there were no gains in wRVUs for APPs across the post-period. However, there was an increase in RVUs over the post-period for physicians, with a 3% increase by day 150 (Supplementary material, Section E). This translates to a predicted increase of approximately 6 RVUs per physician or a $194 increase per physician increase in reimbursement for the week starting day 150, using the 2025 Medicare Physician Fee schedule. Our physician financial productivity metrics were higher that the findings from a recent study that reported ~1.8 more RVUs per week among physicians using an AI scribe.^31^ It is expected that ambient scribe technology may lead to increased billing and risk-adjustment intensity, potentially prompting a re-calibration from the payer side.^32,33^

As previously described, the longitudinal measurements also helped uncover nuanced findings highlighting the adaptation of AI scribe use. For example, a potential advantage of using an AI scribe is that a note is generated based on the active patient-clinician conversation; as a result, a clinician does not require mnemonics or personal notes to recall the details of the encounter when completing the note. This reduces the cognitive load associated with the editing of the note, as the ambient AI scribe-generated transcript reminds the clinician of the encounter details. As clinicians figure out their workflows on “note editing,” given the potentially reduced cognitive effort associated with this task, we observe very interesting patterns. For example, we found that the early post-AI scribe period (first 30 days) was associated with ~5% more notes being closed in the 24 hours after the encounter. However, after day 90, there was no evidence for note closure within 24h compared to the pre-period (Figure 2). This highlights the potential adaptation clinicians made with the AI scribe, recognizing that they did not have to recall––a cognitively challenging task––the details of an encounter when editing a note, instead relying on the completed transcript for recall, which helps in postponing when a note is reviewed and signed.

Similar adaptations in clinician behaviors were also observed from the longitudinal changes in note length and pajama time. As has been reported elsewhere, there have been observed increases in note length^7,9^ and no changes in work outside of work (or pajama time)^13,34^ after AI scribe implementation. This was also observed in our study immediately after AI scribe implementation (day zero); however, as can be seen in Figure 2, over time, there was a declining trend in the note length, with no evidence of differences (compared to the pre-period) in note length by day 150. In contrast, for pajama time there was no evidence of a difference in pajama time till approximately day 90, after which, there was a downward trend, with evidence of time savings by day 150, suggesting that longer-term usage leads to adaptations aligned with the needs of the clinicians.

From a methodological standpoint, there were several innovative aspects. First, as opposed to almost all prior work that has relied on aggregated, clinician-level metrics provided by vendor platforms (e.g., Epic’s Signal), we utilized a patient encounter-level framework. This allowed for the incorporation of important variables (e.g., wRVU, encounter type) for developing homogenous samples. Second, as is evident across the findings, clinicians evolve in their AI scribe use over time. Using single-point measurements (e.g., 1-month before/after or aggregating the data at a single time point) would not capture the true impact of tool adaptations. The current approach allows for characterizing a time-varying effect, reflecting AI scribe adaptation over time. This was also implemented in a robust, interrupted time series framework accounting for serial variability that estimates the treatment effect conservatively through a parsimony enforcing prior. Finally, our encounter-level approach allowed in including all clinicians (physicians, APPs) with AI scribe access over the study period.

This study has several limitations. The study was conducted in primary care settings from 36 clinics that were part of the same health system. Note length was based on the number of characters only; we did not consider the use of smart text or smart phrases (e.g., Epic EHR’s text shortcuts) which influences the number of characters in a note. It is possible that clinicians used more of these in the pre-period given the templated structure of outpatient notes. We used an ITT analysis framework, although there were differences in AI scribe usage in the post-period at the clinician level; the primary assumption was that clinician choice of the AI scribe during an encounter was based on some rational choices. An in-depth study on how scribe tool availability and use in specific types of encounters and clinician and patient characteristics may highlight additional benefits of these tools; in particular, such an analysis would be beneficial to gain a nuanced understanding of the differential benefits of AI scribe use among clinicians treating more complex patient populations. Additional analysis including patient complexity, disease burden, level of service and related aspects may provide insights into how AI scribes are potentially beneficial and various patient care scenarios. We used an interrupted time series approach to estimate the effect associated with AI scribe tool introduction. This analysis is dependent upon the assumption that typical serial variability will follow a consistent pattern across the study period, which appears reasonable. However, an approach where physicians are selected into treated and control groups may offer a more robust approach to estimating treatment effects through the use of explicitly defined contemporaneous randomized treatment and control groups. Finally, the current study did not gather clinician perspectives on AI scribe use using qualitative methods, which might provide insights into the clinician adaptation processes (e.g., why the time savings changed over time).

## Data Availability

Data used for this study is not publicly available.

## Funding

The funding for this study was provided in part by the Center for Health AI (a joint Washington University School of Medicine and BJC Health System endeavor), the Professional Fulfillment Program (WashU Division of General Internal Medicine) and the Barnes-Jewish Hospital Medical Staff Association.

## Conflict of Interests

Outside of this work, Dr. Kannampallil serves as an editor for an Elsevier journal and Dr. Waken serves as a statistical editor for another Elsevier journal. The other authors report no conflicts of interest. Dr. Kannampallil is an unpaid advisor on the research steering committee for Abridge. Dr. Maddox is a paid advisor on FDA’s Digital Health Advisory committee, an advisory board member for Elion and a board director for the J.F. Maddox Foundation.

## Contributor Statement

TK, RJW, and SSL conceived the study. TK and RJW were involved in the data analysis. All authors were involved in the interpretation of the results, writing of the manuscript and provided approval for the final version of the manuscript.

## Data Availability

The data underlying this study cannot be shared publicly as it contains identifying information that cannot be removed without compromising the quality of the shared data.

## Figure Legend

**Figure 1.** STROBE diagram (within inclusion and exclusion criteria)

**Figure 2.** IRRs over in the post AI scribe period from day zero to day 168 (i.e., 24 weeks, post-period) for each of the considered outcomes. Note: in the manuscript we report IRRs at specific time intervals of 0, 30, 60, 90, 120 and 150 days corresponding to approximately monthly intervals

## Notes

### Competing Interest Statement

Thomas Kannampallil is an unpaid advisor for Abridge Inc.

### Summary of Updates

changed text to update based on reviewer comments

