## Supplement for "Longitudinal effects of ambient AI scribe use on documentation burden and financial productivity in primary care: A quasi-experimental study"

Supplementary Materials

### Section A: Description of the outcome variables

**Appendix Table 1.** Detailed description of the outcomes of interest. Note: Work outside of Work (WOW) was the aggregate of TOSH and pajama time.

| **Measure** | **Definition** |
| --- | --- |
| Note writing time (primary) | This was measures using *active time* spent on writing notes. This included recording of active keystrokes, mouse movements or scrolling. |
| Time outside of scheduled hours writing notes (TOSH) | Time spent on documentation outside of patient encounters on days with scheduled patient appointments |
| Pajama time spent on writing notes | Time spent on documentation outside of 7:00 am to 5:30 pm |
| Note length | Total number of characters associated with a finalized note. |
| Note closure within 24 hours | Time elapsed from the scheduled end of the patient encounter to when the encounter is deemed closed; classified as a binary variable (<24h vs. >24h) |
| Work Relative Value Units (wRVU) | A standardized measure of a clinician’s work effort for reimbursement. This was retrieved from billing data representing billed wRVU for an encounter. |

### Section B: Model description

#### Spline specification

Our thin plate spline generalized additive model specification has twelve knots, chosen such that we estimate, at most, one interruption effect regression coefficient parameter per two weeks in the interruption period. Our choice of the number of knots was based on preliminary, exploratory analyses using Poisson generalized additive models that focused on prioritizing parsimony, model fit, consistency in modeling choices across all outcomes, and interpretability.

#### Modeling specification

##### Functional form

We specified a non-linear intervention effect starting at day zero (i.e., day of AI scribe access) characterized by an intercept shift as well as a thin plate spline. Specifically, we used the following model:

$$E\left[ y_{i} \right]=f^{-1}\left( \mu\left( t_{i} \right)+\alpha+\gamma_{i}+g\left( t_{i}, \beta\right) \right)$$

where $f\left( \cdot\right)$ is the natural log, $i$is the observation index for each encounter, $t_{i}$ is the day index for encounter $i$, $\mu\left( t_{i} \right)$ is a latent time series component that is a function of the day index $t_{i}$, $\alpha$ is a time invariant mean term, $\gamma_{i}$ accounts for overdispersion, and $g\left( t_{i}, \beta\right)$ is our non-linear interruption effect.

##### Prior specification choices

We incorporate a regularized horseshoe prior specification^1^ (Piironen and Vehtari) on the interruption effect of interest to encourage parsimony and discourage the model from attributing typical post treatment variability to an interruption effect, and a penalized complexity prior^2^ (Simpson) to aid in partitioning serial and extra-Poisson variability in our count data analyses. We use default prior choices for both the thin plate spline penalty terms ($\lambda_{1}, \lambda_{2}$) as well as the autoregressive mean processes as are implemented in the mvgam package.^3^

##### Full model specification

###### Shared components

In both models, we specify

$$\boldsymbol{\alpha} \sim N\left( \boldsymbol{\alpha}_{\boldsymbol{0}}, 2 \right),$$

$$\boldsymbol{\lambda} \sim N^{+}\left( 5, 30 \right),$$

$$\tau\sim C^{+}\left( 0, 1 \right),$$

$$\psi\sim gamma\left( 1, 1 \right),$$

$$\beta_{0} \sim N\left( 0, \tau+c+\psi\right),$$

$$\boldsymbol{\beta}_{-0} \sim N\left( \boldsymbol{0}, \left( \tau+c+\psi\right)^{2}\left( \lambda_{1}\boldsymbol{S}_{1}+ \lambda_{2}\boldsymbol{S}_{2} \right)^{-1} \right)$$

$$\nu\sim N^{\left( -1, 1 \right)}\left( 0, \nu_{0} \right),$$

$$\rho\sim N\left( 0, \rho_{0} \right),$$

$$\sigma\sim N^{+}\left( 0, \sigma_{0} \right),$$

$$\mu_{1}\sim N\left( 0,\sigma\sqrt{\rho/w} \right),$$

$$\mu_{i\in2:n} \sim N\left( \nu\mu_{i-1}, \sigma\sqrt{\rho/w} \right),$$

$$\phi_{i} \sim N\left( 0, \sigma\sqrt{1-\rho} \right),$$

where $\boldsymbol{\beta}_{-0}$ is the spline coefficient parameter vector with no intercept, $\beta_{0}$ is the intercept shift at treatment time zero, $c$ is a constant term set to 0.1 as part of the regularized horseshoe specification, $\boldsymbol{\alpha}_{\boldsymbol{0}}$ is a user input vector where the first value is the pre-period average of the process and all other values are set to zero, $\boldsymbol{S}_{k}$ are penalty matrices for our thin plate spline, $\sigma_{0}$ and $\rho_{0}$ are user supplied input values (set at 0.5 and 1 respectively), and $w$ is a scaling factor in the penalized complexity framework that helps us efficiently partition variability between the overdispersion component $\phi_{i}$ and the latent time series process component $\mu_{i}$ without the need for strong priors, and superscripts on distributions indicate the support for the parameter of interest ($+$ indicates the set of positive real numbers).

##### Likelihood specification

###### Count data model specific components

For all countable outcomes, such as time-based outcomes, number of note closures within 24 hours of visit, as well as note length in characters, observations were characterized as counts (seconds for time based outcomes) per encounter. In our count data processes (all non-RVU outcomes), we further specify $\nu_{0}=0.5$ and

$$y_{i} \sim Poisson\left( \boldsymbol{x}_{i}\boldsymbol{\alpha}+g\left( t_{i}, \boldsymbol{\beta} \right)+\mu_{i}+\phi_{i} \right).$$

###### RVU model specific components

Our RVU model is specified in a way that uses the same overdispersion framework (which is equally flexible) as the Poisson specification instead of taking advantage of the additional parameter in the likelihood, which we set to one, which makes this equivalent to an exponential distribution. We do use the log link function in spite of this not being the canonical link function in the context of a generalized linear model parameterization. Here, we set $\nu_{0}=0.5$ and specify

$$y_{i} \sim gamma\left( 1,\exp\left( -\boldsymbol{x}_{i}\boldsymbol{\alpha}-g\left( t_{i}, \boldsymbol{\beta} \right)-\mu_{i}-\phi_{i} \right) \right).$$

#### Model Diagnostics

In all simulations, we defined posterior convergence when all $\hat{R}$ values were less than 1.05 and visually inspected traceplots for all parameters of interest appeared to converge to a single independent process across all chains.

#### Use of sufficient statistics to facilitate computationally efficient model fitting

Instead of fitting our model to all encounters as individual observations, we take advantage of the fact that the sum of Poisson count data processes also has a Poisson distribution; our outcome is then the sum of responses indexed to treatment day, and we include an offset term representing the number of encounters included in each aggregated sum. In addition to reducing computational burden, this also allows us to simplify our modeling strategy when investigating note closures within 24 or after 72 hours, as we can then treat those like count data processes as well.

We use a similar strategy to model the sum of gamma processes, as for identically distributed $x_{i} \sim gamma(\alpha, \beta)$, $\sum_{i=1}^{n} x_{i} \sim gamma\left( n\alpha, \beta\right).$

#### Predictions for a new physician week

We generate predictions for new physician weeks starting at days 0, 30, 60, 90, 120, and 150 using the modeled MCMC draws for all parameters, the likelihood specified, as well as the assumptions that physicians work 5 of 7 days per week, and that the average number of encounters per physician day is constant across the time period (at 19.34 encounters per day). We calculate the weekly outcomes under both the observed and counterfactual processes as well as their difference a) by randomly generating each of the outcomes across 19.34 encounters (specified using an offset term) in each day in the week in question, finding the mean, 2.5^th^ and 97.5^th^ percentiles of the generated MCMC draws, and directly report these summaries difference in observed and counterfactual process draws for inferences.

### Section C: Time series plots

The figure below gives estimates of a typical encounter averaged at the day level for each of the response processes.

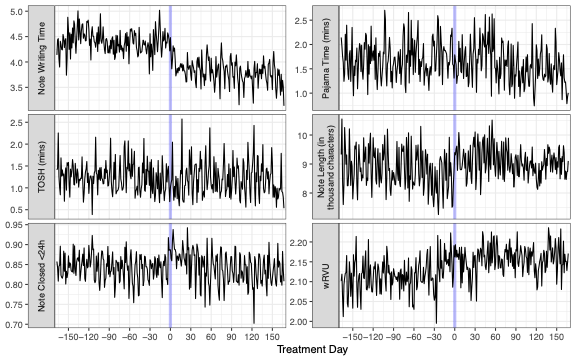

### The plot below gives the number of encounters by day.

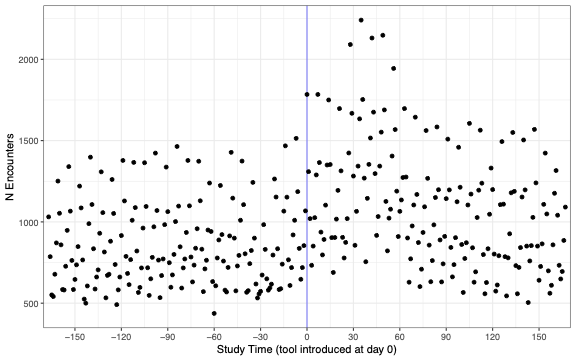

### Section D: IRRs and predicted changes at additional time points

The following tables provide a summary of the IRRs for each of the outcome variables at days 30, 60, 90, and 120 (reflecting Figure 2 in the main manuscript) and the predicted changes for week starting the corresponding day.

| **Outcome measure** | **IRR (95% CI) at *Day 30*** | **Predicted Change at *Day 30*** |
| --- | --- | --- |
| Note writing time (mins) | 0.88, (0.86, 0.91)* | 50.80, (33.96, 68.74)* |
| Pajama time (mins) | 1.00, (0.91, 1.11) | -0.95, (-28.67, 27.37) |
| Time Outside of Scheduled Hours (mins) | 0.95, (0.85, 1.06) | 6.08, (-17.80, 30.13) |
| Note length (1000 Characters) | 1.04, (1.01, 1.07)* | 0.01, (-3.82, 4.05) |
| Note closed <24h | 1.02, (1.00, 1.04) | 1.43, (-20.00, 22.14) |
| Total work Relative Value Units (wRVU) | 1.02, (1.01, 1.03)* | 2.76, (-44.04, 53.08) |

| **Outcome measure** | **IRR (95% CI) at *Day 60*** | **Predicted Change at *Day 60*** |
| --- | --- | --- |
| Note writing time (mins) | 0.87, (0.85, 0.90)* | 52.86, (36.40, 70.23)* |
| Pajama time (mins) | 0.98, (0.89, 1.08) | 4.43, (-18.20, 28.88) |
| Time Outside of Scheduled Hours (mins) | 0.97, (0.88, 1.08) | 3.03, (-18.66, 26.44) |
| Note length (1000 Characters) | 1.04, (1.01, 1.07)* | 0.00, (-4.03, 4.09) |
| Note closed <24h | 1.00, (0.98, 1.01) | -0.71, (-21.43, 20.00) |
| Total work Relative Value Units (wRVU) | 1.03, (1.02, 1.04)* | 3.86, (-45.93, 52.18) |

| **Outcome measure** | **IRR (95% CI) at *Day 90*** | **Predicted Change at *Day 90*** |
| --- | --- | --- |
| Note writing time (mins) | 0.86, (0.84, 0.88)* | 60.49, (44.73, 77.92)* |
| Pajama time (mins) | 0.88, (0.80, 0.97)* | 20.25, (-5.68, 46.35) |
| Time Outside of Scheduled Hours (mins) | 0.97, (0.87, 1.07) | 4.15, (-20.69, 28.87) |
| Note length (1000 Characters) | 1.02, (0.99, 1.05) | -0.12, (-3.71, 3.89) |
| Note closed <24h | 0.99, (0.97, 1.01) | -0.71, (-22.14, 22.14) |
| Total work Relative Value Units (wRVU) | 1.02, (1.01, 1.04)* | 4.90, (-46.30, 53.23) |

| **Outcome measure** | **IRR (95% CI) at *Day 120*** | **Predicted Change at *Day 120*** |
| --- | --- | --- |
| Note writing time (mins) | 0.85, (0.83, 0.87)* | 61.43, (45.89, 78.49)* |
| Pajama time (mins) | 0.86, (0.79, 0.94)* | 26.32, (-2.68, 54.49) |
| Time Outside of Scheduled Hours (mins) | 0.95, (0.85, 1.07) | 6.82, (-19.56, 35.31) |
| Note length (1000 Characters) | 1.01, (0.98, 1.04) | -0.05, (-4.02, 3.86) |
| Note closed <24h | 0.99, (0.97, 1.00) | -0.71, (-21.43, 19.3) |
| Total work Relative Value Units (wRVU) | 1.02, (1.01, 1.03)* | 4.82, (-42.12, 52.73) |

### Section E: Sensitivity analyses

#### Clinician age group and sex adjusted results

We implement the modeling process described while incorporating time invariant trends for physician age group (<36, 36-55, >55) at first included encounter and sex as predictors in $\boldsymbol{x}_{i}$ to account for potential changes in provider makeup throughout the study period. If we look at our incidence rate ratios that represent the average effect by response, we see very similar trends to those produced in Figure 2 in the main body of the paper.

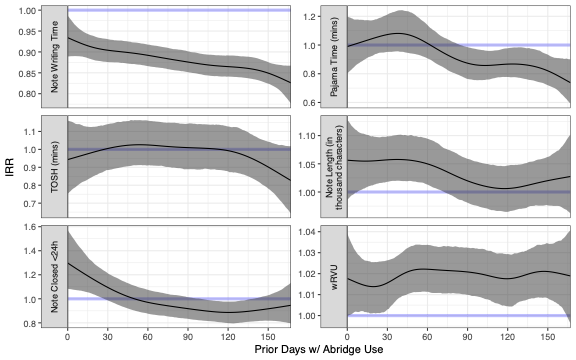

#### Stratified results by clinician type

As mentioned in Table 1, our encounter sample consisted of 90 physician (MD/DO) and 124 non-physician (APP) providers; the physician providers make up 76.5% of all encounters included in our analysis. To investigate whether the Ambient AI scribe tool affected these groups differentially, we stratified our sample into each provider type, performed all described analyses, and report the typical effect (as measured by incidence rate ratios) over time for each of the outcomes below.

Our results within the physician stratum look very similar to the overall results, which is unsurprising given that they make up the majority of our sample; we do see larger overall reductions by day 150 in active minutes, pajama time, but smaller immediate shifts on average. Most notably, RVUs appear to slightly increase over time on average within the physician group. In the APP group, we do not see the same consistent trend downward in active minutes or PJ time, and see no evidence of an effect on average in RVUs than in the pre-period.

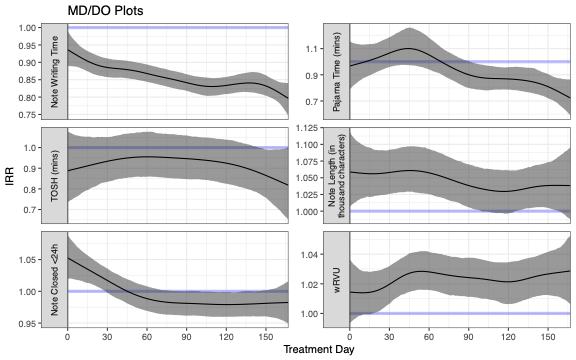

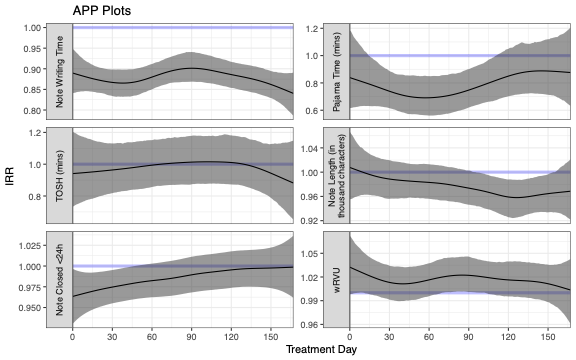
